# Clinical Deep Phenotyping of Treatment Response in Schizophrenia (CDP-STAR): design and methodology of a prospective multimodal observational study

**DOI:** 10.1101/2025.06.17.25329786

**Authors:** Vladislav Yakimov, Lara Neuwinger, Madeleine M. Weber, Maximilian Brantl, Isabel Maurus, Jana Sautner, Miriam John, Berkhan Karslı, Genc Hasanaj, Anne Bungard, CDP-Working Group, Alkomiet Hasan, Elias Wagner, Laura Fischer, Paula Steiner, Benedikt Schworm, Siegfried Priglinger, Sergi Papiol, Peter Falkai, Andrea Schmitt, Florian J. Raabe, Daniel Keeser, Lukas Roell, Joanna Moussiopoulou, Emanuel Boudriot

## Abstract

Schizophrenia spectrum disorders (SSDs) exhibit a marked heterogeneity in clinical course and treatment outcomes. Some individuals achieve remission and recovery, whereas others experience repeated relapses and progressive deterioration in psychosocial functioning. This variability underscores the unmet clinical need for prognostic biomarkers to predict treatment outcomes and guide personalized care. Deep phenotyping with multimodal data integration holds promise for understanding this complexity and delivering clinically relevant predictive models of treatment response in SSDs. To address this need, we initiated the Clinical Deep Phenotyping of Treatment Response in Schizophrenia (CDP-STAR) study, a prospective, naturalistic, longitudinal observational study integrating comprehensive multimodal assessments. These include clinical phenotyping, magnetic resonance imaging (MRI), electroencephalography (EEG), retinal imaging, and extensive sampling of blood and cerebrospinal fluid (CSF) for multi-omics profiling. The study aims to externally validate promising biomarker candidates and elucidate the pathophysiological mechanisms underlying treatment outcomes. This innovative deep phenotyping framework integrates data across multiple critical domains, enabling external validation of potential biomarkers and the discovery of novel ones. Ultimately, the CDP-STAR study aims to yield mechanistic insights that advance precision psychiatry and inform clinical decision-making in SSDs.

## 1. Introduction

Schizophrenia spectrum disorders (SSDs) are characterized by considerable clinical heterogeneity and highly diverse disease trajectories. While some individuals achieve remission or even recovery, others experience recurrent relapses and progressive functional decline [1]. This clinical variability is accompanied by significant neurobiological heterogeneity, as evidenced by increased brain structural variability [2],[3] and a highly polygenetic architecture [4]. Such heterogeneity in biological underpinnings has been suggested to contribute to the interindividual variability in pharmacological treatment resistance, which occurs in up to one-third of affected individuals.

To date, psychiatry still lacks reliable prognostic biomarkers to inform clinical decision-making [7]. A personalized medicine approach based on such markers holds great promise for improving patient care and well-being, as well as alleviating socioeconomic burden [8]. For instance, the early recognition and stratification of individuals with treatment-resistant schizophrenia (TRS) could minimize ineffective treatment trials, facilitate the timely initiation of adequate treatments such as clozapine, and enable more efficient allocation of clinical resources.

SSDs are associated with widespread alterations across various biological systems[9],[10],[11],[12], including changes in brain structure [13], microstructure [14], and connectivity [15] in multiple neuronal networks [16],[17]. Among these, the striatum has emerged as a central hub, repeatedly implicated in the pathophysiology of SSDs [18],[19]. This is particularly relevant given that most antipsychotics exert their effects primarily through dopamine D_2_ receptor blockade in the striatum [18]. The functional striatal abnormalities (FSA) score has been internally and externally validated as a diagnostic biomarker, successfully differentiating individuals with schizophrenia from healthy controls (HC) with a balanced accuracy exceeding 80% in two independent studies [18],[20]. The FSA score has also been associated with poor short-term treatment response [18], although these findings warrant further replication. Another striatal biomarker candidate, the striatal connectivity index (SCI), has shown potential in differentiating responders from non-responders and predicting relapse risk [21],[7],[22]. Beyond the striatum, additional prognostic biomarkers have been proposed, including functional connectivity between bilateral superior temporal cortex and other cortical regions [23], glutamate levels in the anterior cingulate cortex measured via magnetic resonance spectroscopy [24], and neuromelanin-sensitive MRI signal [25]. In the domain of cognitive dysfunction, the electroencephalography (EEG)-based mismatch negativity (MMN) has also emerged as a promising biomarker candidate due to its robust test-retest reliability [26],[27].

Although numerous prognostic biomarker candidates have been proposed, the majority of evidence to date is limited to in-sample statistical associations [7]. While some markers have undergone internal validation, external validation in naturalistic samples – a crucial prerequisite for clinical translation [28] – remains scarce [29]. Moreover, most candidate biomarkers focus on single parameters or systems and were derived from cross-sectional studies, limiting their capacity to capture the heterogeneity and complexity of SSDs and reducing their translational potential. A deeper understanding of the biological mechanisms underlying clinical outcomes, such as treatment response or treatment resistance, could refine existing biomarkers and support the discovery of novel candidates in a mechanistically informed manner.

A longitudinal, naturalistic, and multimodal research framework is warranted to advance our understanding of the underlying biology and enable the validation of biomarker candidates [29] as well as the discovery of novel ones, thereby fostering progress toward precision medicine in psychiatry. To this end, we initiated the Clinical Deep Phenotyping of Treatment Response in Schizophrenia (CDP-STAR) study, a prospective, naturalistic, observational study employing a comprehensive multimodal approach. The overarching goal of this translational project is to evaluate the predictive capability of prognostic biomarker candidates and elucidate the pathophysiological mechanisms underlying outcome trajectories in SSDs. Based on a previous study [30], our protocol integrates detailed clinical phenotyping with multimodal neuroimaging, EEG, retinal assessments, and multi-omics profiling of blood and cerebrospinal fluid. This design enables both the external validation of existing biomarker candidates and the investigation of short- and long-term trajectories of SSDs across genetic, molecular, cellular, and systems biology levels. Ultimately, this multidimensional framework aims to generate mechanistic insights into the complex pathophysiology of SSDs and advance the development of precision psychiatry.

## 2. Methods and design

The CDP-STAR study is a naturalistic, multimodal, prospective, longitudinal, single-center study conducted at the Department of Psychiatry and Psychotherapy, University Hospital of the Ludwig-Maximilian University Munich, Germany. It is an add-on study to the *Munich Mental Health Biobank* (project number 18-716) [31], and was approved by the local ethics committee (project number 24-0341, dated 08.07.2024). The study is registered at the German Clinical Trials Register (DRKS00034820) and OSF (https://doi.org/10.17605/OSF.IO/SQ2TZ).

The multimodal protocol of the longitudinal CDP-STAR study builds upon the previous Clinical Deep Phenotyping (CDP) study [30]. From 09.10.2020 to 10.11.2023, we collected cross-sectional data from 466 participants, highlighting the feasibility of our approach.

### 2.1. Study population

This study includes inpatients with SSDs according to the Diagnostic and Statistical Manual of Mental Disorders 5th Edition, text revision (DSM-5-TR, Version 7.0.2). The diagnoses are validated with the Mini-International Neuropsychiatric Interview (M.I.N.I.) [32]. Further inclusion criteria include an age between 18 and 65 years, hospitalization due to newly developed or exacerbated psychotic symptoms, ability to provide informed consent, and fluency in the German language. Exclusion criteria include a primary psychiatric disorder other than SSDs, pregnancy, patients with acute danger to self and/or others (e.g., acute suicidality), individuals who are not able to provide informed consent, currently undergo coercive treatment or involuntary hospitalization at the time of study inclusion, individuals with relevant neurological comorbidity (e.g., dementia, multiple sclerosis, epilepsy) as assessed by experienced clinicians or psychotic symptoms due to a general medical condition.

Inpatients in our clinic are screened by a study physician for inclusion and exclusion criteria on a regular basis, and written informed consent is obtained prior to any study-related procedures.

### 2.2. Study timeline

The study flowchart is depicted in Figure 1. After study inclusion, multimodal assessments, including multimodal magnetic resonance imaging (MRI), electroencephalography (EEG), retinal examinations, clinical characterization, blood sampling, and, if clinically indicated, cerebrospinal fluid (CSF) sampling, are being performed at baseline (V1) and four weeks after inclusion (V2). Clinical assessments are repeated three months (V3), six months (V4), and two years (V5) after inclusion to characterize the participants regarding short- and long-term disease outcomes. Besides patient interview, therapeutic drug monitoring is conducted at baseline (V1) to assess pharmacological treatment adherence.

**Figure 1.**
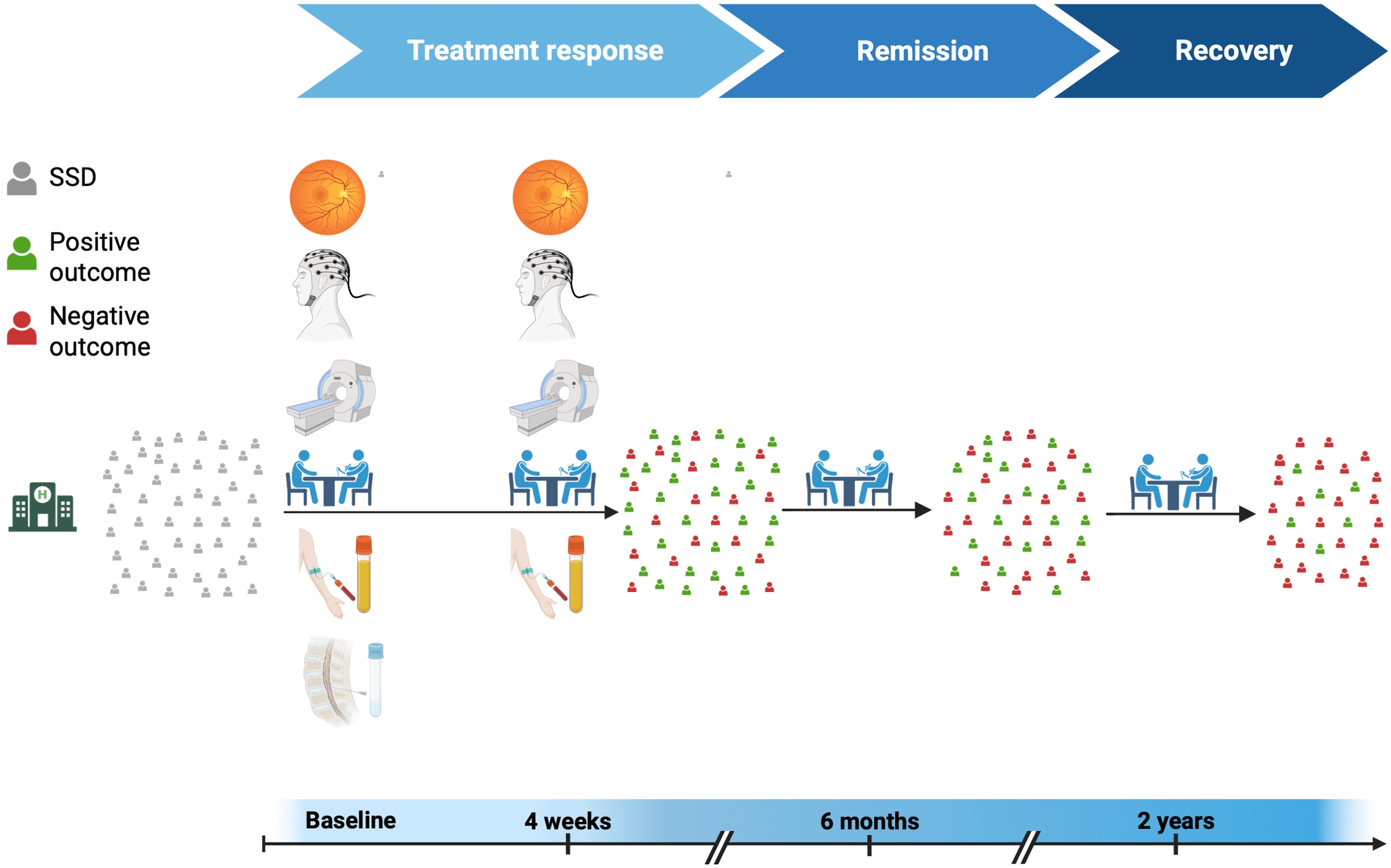
Study design and timeline. Abbreviations: SSD, schizophrenia spectrum disorders. Created in BioRender. Yakimov, V. (2025).

### 2.3. Medical examinations

#### 2.3.1. Clinical characterization

Table 1 provides an overview of the examinations performed during the study visits. The clinical characterization includes diagnosis validation and assessment of current and past suicidality using the Mini-International Neuropsychiatric Interview (M.I.N.I.) [32] according to DSM-5. The CDP-STAR assessments comprise the basic phenotyping framework of the *Munich Mental Health Biobank*[31], which includes (A) a structured assessment of socioeconomic status, psychiatric and medical history, as well as family history of mental disorders, and (B) a set of transdiagnostic self-ratings. These self-ratings have been described previously by our group[30] and include the Childhood Trauma Screener (CTQ-Screen) [33], the Brief Resilience Scale [34], the Loneliness Scale [35], the Lubben Social Network Scale [36], the World Health Organization-5 Well-Being Index (WHO-5) [37], the abbreviated version of the World Health Organization Quality of Life Scale (WHOQOL-BREF) [38], the Patient Health Questionnaire-9 (PHQ-9) [39], and the Munich Chronotype Questionnaire (MCTQ) [40].

**Table 1.**
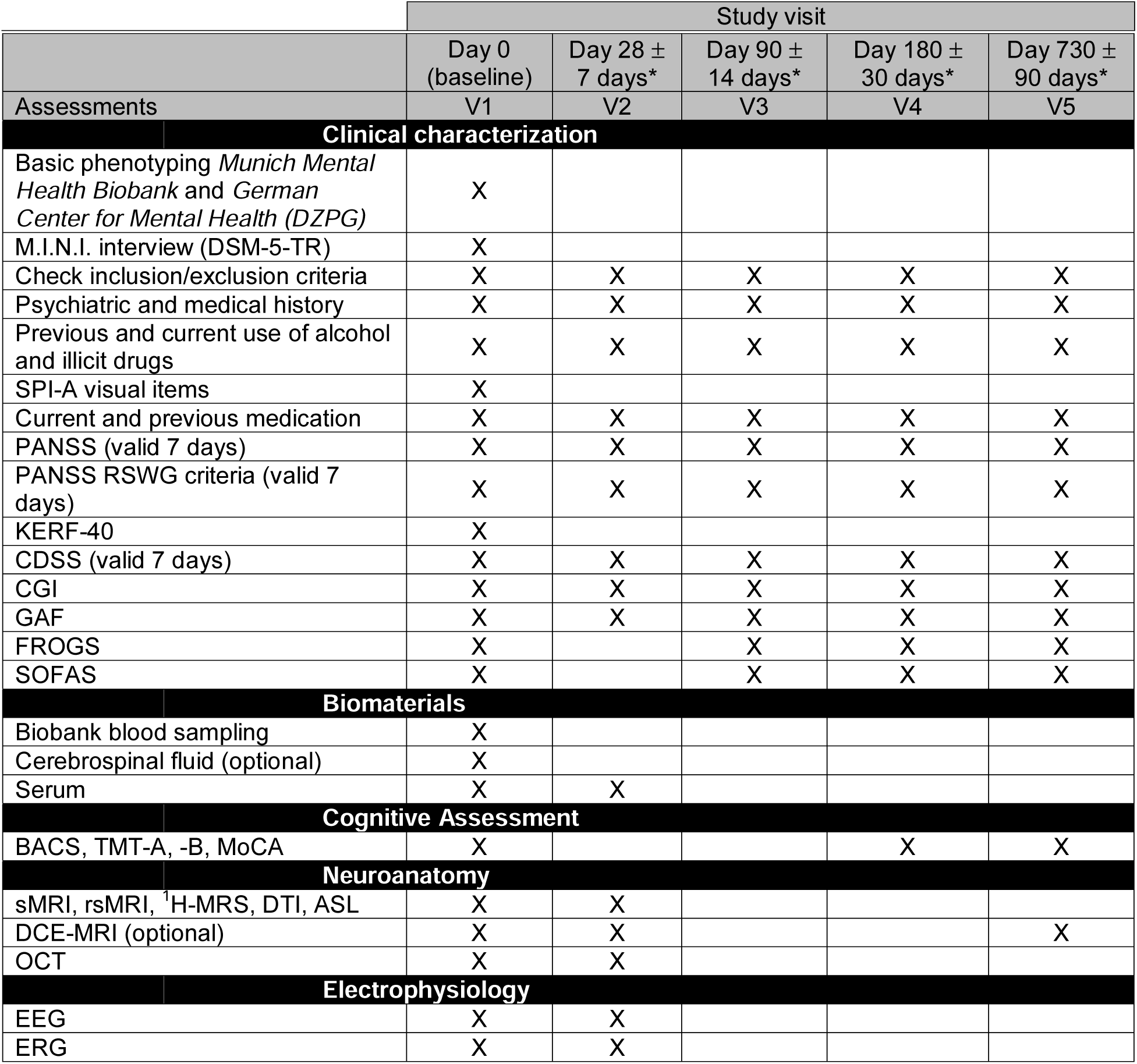
Overview of baseline and follow-up examinations. The basic phenotyping of the *Munich Mental Health Biobank* has been described in detail by Kalman et al., 2022 [31] and Krčmář et al., 2023 [30]. Abbreviations: ASL, Arterial Spin Labeling; BACS, Brief Assessment of Cognitive Function in Schizophrenia; CGI, Clinical Global Impression; CDSS, the Calgary Depression Scale for Schizophrenia; CTQ-S, Childhood Trauma Questionnaire screener; DCE, dynamic contrast-enhanced; DTI, Diffusion Tensor Imaging; EEG: electroencephalography, ERG: electroretinography; FROGS, Functional Remission of General Schizophrenia; GAF, Global Assessment of Functioning; ^1^H-MRS, Magnetic Resonance Spectroscopy; MoCA, Montreal Cognitive Assessment; OCT: optical coherence tomography, PANSS, Positive and Negative Syndrome Scale; rsfMRI, resting-state functional MRI; sMRI, structural MRI; SOFAS, Social and Occupational Functioning Assessment Scale; TMT, Trail Making Test. *Indicates working days.

The study-specific clinical phenotyping includes the Positive and Negative Syndrome Scale (PANSS) [41], the Calgary Depression Rating Scale for Schizophrenia (CDSS) [42], the Clinical Global Impression (CGI) scale [43], the Global Assessment of Functioning (GAF) scale [44], the Functional Remission of General Schizophrenia (FROGS) scale [45], the Social and Occupational Functioning Assessment Scale (SOFAS) [46], the visual items of the Schizophrenia Proneness Instrument – Adult Scale (SPI-A) [47], as well as a questionnaire covering adverse childhood experiences (“Belastende Kindheitserfahrungen”, KERF-40) [48]. Neurocognitive functioning is assessed via the standardized Brief Assessment of Cognition in Schizophrenia (BACS) [49]. This battery (30 – 45 min) assesses six cognitive domains identified as important for clinical trials in schizophrenia by the MATRICS Neurocognition Committee [50]. These include verbal memory, working memory, motor speed, attention, executive functions, and verbal fluency [49]. In addition, we complement the battery with the time-efficient and widely used Montreal Cognitive Assessment (MoCA) [51] and Trail Making Test (TMT): Parts A & B [52] (Table 1). The battery of assessments is performed by trained mental health professionals.

Furthermore, a comprehensive medical and psychiatric history, as well as treatment data (e.g., medication, electroconvulsive therapy), are collected through patient interview and medical chart review (Table 1). This includes the duration of illness, age of symptom onset, duration of untreated psychosis, number of hospitalizations due to psychosis, hearing impairment during the past twelve months, lifetime cannabis use, and substance abuse during the past seven days. Following our previous protocol, we assess past and present medical conditions, including neurological disorders, cardiometabolic comorbidities, risk factors (e.g., body mass index, smoking status, dyslipidemia), and ophthalmological conditions. Electronic health records are endorsed to verify the collected data if applicable and available. These comprehensive clinical data enable us to estimate the environmental risk score for schizophrenia [53] and the future risk of cardiovascular disease [54] in our cohort.

#### 2.3.2. Multimodal brain imaging

Multimodal Magnetic Resonance Imaging (mMRI) is performed with a Siemens Magnetom Prisma 3T MRI scanner (Siemens Healthineers, Erlangen, Germany). It includes anatomical (T1-weighted magnetization prepared-rapid acquisition gradient echo [T1-MPRAGE], T2 sampling perfection with application-optimized contrasts using different flip angle evolution [T2-SPACE], diffusion tensor imaging [DTI]) and functional (resting-state functional MRI, multivoxel magnetic resonance spectroscopy [MRS] of the anterior cingulate cortex, dynamic contrast-enhanced MRI [DCE-MRI], and arterial spin labeling [ASL]) measurements. For the mMRI measurements, the Human Connectome Project (HCP) protocol [55] is used. For the DCE-MRI measurement, a gadolinium-based contrast agent (Gadobutrol, Gadovist®, Bayer AG, Leverkusen, Germany) is administered intravenously via the antecubital vein at a dose of 0.1 mmol/kg. The injection is delivered at a rate of 3 mL/s, followed by a 25–30 mL saline flush.

#### 2.3.3. Electroencephalography (EEG)

Digitalized resting-state EEG recordings are performed with a standardized set-up (BrainAmp amplifier, Brain Products, Martinsried, Germany), including 32 scalp electrodes (10/20 system). All EEGs are recorded during wakeful rest under two conditions: (i) eyes closed and (ii) eyes open while fixating on a central point.

#### 2.3.4. Retinal anatomy and electrophysiology

Due to the neurodevelopmental origin of the retina[56] and emerging evidence for alterations in individuals with SSDs [57],[58],[10],[59], our phenotyping includes the investigation of retinal anatomy by optical coherence tomography (OCT) (ZEISS CIRRUS HD-OCT 5000 device, Carl Zeiss Meditec AG) and retinal electrophysiology by electroretinography (ERG) (RETeval electroretinography, LKC Technologies, Inc.) [30]. In addition, intraocular pressure, refraction, and best-corrected visual acuity are measured in all participants at baseline.

#### 2.3.5. Biobanking of blood and cerebrospinal fluid

The biobanking of samples is provided by the *Munich Mental Health Biobank* [31] infrastructure, as previously described [30]. Blood-based biobanking includes the following: 1 x 7,5 ml K3EDTA tube (Fa Sarstedt, Cat no 01.1605.001) for DNA extraction, 1 x S-Monovette® RNA exact (Fa Sarstedt, Cat no 01.2048.001) for RNA extraction, 1 x 9 ml K3EDTA tube (Fa Sarstedt, Cat no 02.1066.001) for plasma-based analysis, and 1 x 9 ml tube with coagulation activator (Fa Sarstedt, Cat no 02.1063.001) for serum-based analysis. All samples are stored at –80°C after initial processing. Besides blood samples, cerebrospinal fluid (CSF) is also collected and stored at –80°C from individuals with SSDs who consent and undergo a diagnostic lumbar puncture.

### 2.4. Outcome measures

The primary outcome (O1) of this study is the evaluation of prognostic capabilities (measured by the area under the curve (AUC) in receiver operator characteristic (ROC) curves) of biomarker candidates (e.g., FSA score [18]) regarding acute treatment response in patients with SSDs four weeks after inclusion (Figure 2). Short-term treatment response would be defined as ≥25% symptom reduction in PANSS total score compared to the baseline score [28]. To ensure our results are not driven by idiosyncrasies in defining treatment response, we will include two other definitions commonly used in the field: ≥50% symptom reduction in the PANSS total score and percentage change with baseline correction as previously described [28].

**Figure 2.**
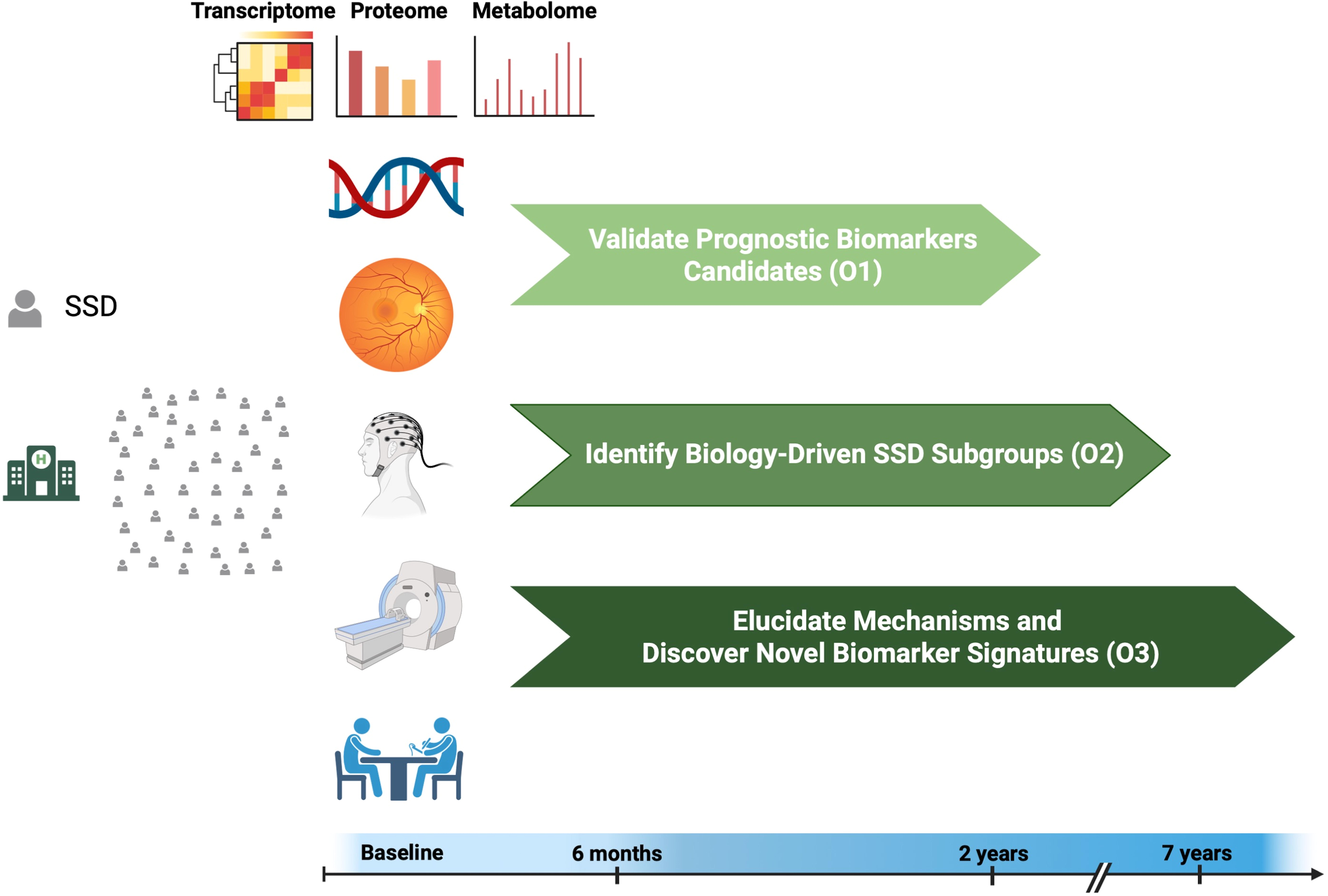
Study objectives. Abbreviations: O1, objective 1; O2, objective 2; O3, objective 3; SSD, schizophrenia spectrum disorders. Created in BioRender. Yakimov, V. (2025).

Secondary outcomes include investigating the prognostic capabilities of candidate biomarkers regarding treatment resistance (at least two unsuccessful adequate trials of antipsychotics) [60], relapse [61], symptomatic remission six months after inclusion [62], and recovery (defined as improvements of both clinical and social domains) [63] two years after inclusion in SSDs. The definition of symptomatic remission will be based on modified Andreasen remission criteria [62] six months after baseline measurements. We will also aim to identify biologically driven subgroups within the schizophrenia spectrum (secondary outcome) (O2) using unsupervised machine learning models in our multimodal data. Further secondary outcomes include exploring the neurobiological and systemic mechanisms behind acute treatment response, treatment resistance, relapse, symptomatic remission, and recovery (O3), employing a multimodal approach (Figure 2).

### 2.5. Power analysis and expected sample size

In the context of our ROC analysis, we conducted a power calculation to determine the predictive capability of candidate biomarkers. The expected Area Under the Curve (AUC) was set at 0.7, with an aim to achieve a power of 80% at a significance level (α) of 1%. Since there is limited data regarding the predictive capabilities of candidate biomarkers in terms of treatment response, we set the expected AUC to 0.7 for two reasons. First, we expect the AUC of the FSA score (as the most advanced biomarker in schizophrenia [7]) to be lower for outcomes such as treatment response than its AUC for discriminating schizophrenia from healthy controls (0.8) [18],[20]. Second, it has been suggested that for a biomarker candidate to achieve clinical utility, it should show an AUC of at least 0.7 [8]. Considering the ratio of non-responders to responders in SSDs, approximated at 1.0 [64], the required sample size was estimated. The calculation was performed using the R programming language with the *power.roc.test* function from the pROC package[65]. The analysis indicated the need for at least 132 participants (66 responders and 66 non-responders) to attain the desired statistical power for our ROC analysis.

Based on our power analysis, we would need at least 132 participants to attain enough statistical power for our primary outcome analysis. We assume a cumulative drop-out rate of approximately 25% during follow-up (4 weeks after inclusion) from previous studies [66],[67]. Furthermore, based on our previous multimodal Clinical Deep Phenotyping (CDP) study [30], we estimate that approximately 50% of the participants would yield complete datasets (including clinical phenotyping, MRI, EEG, retinal investigations, and blood analyses at baseline and V2), which would be necessary for our primary outcome. Taking the expected drop-out rate and partial data missingness into account, we would need to recruit 352 participants with SSDs to yield the necessary 132 patients for the primary analysis as follows: 352*0.75 [minus drop-out] *0.5 [minus participants with incomplete dataset] = 132 participants.

For secondary outcomes such as identifying biological subgroups within the schizophrenia spectrum, we would pool cross-sectional and baseline data from the previous CDP study (n = 233 individuals with SSDs), from the ongoing interventional BrainTrain study (expected n = 120 individuals with SSDs), and the current CDP-STAR study (n = 352 participants), which have harmonized protocols for baseline measurements. Based on our previous CDP study [30], we expect complete baseline/cross-sectional datasets in approximately 70% of the participants, yielding a total of 494 participants: (233+120+352)*0.7 [minus participants with incomplete dataset].

## 3. Study progress

As of 12.06.2025, we have recruited 44 individuals with schizophrenia spectrum disorders. Of those, 36 have complete data (incl. clinical data, MRI, EEG, retinal imaging data, and blood samples) at baseline. So far, 31 out of 41 individuals have participated in V2, resulting in a current dropout rate of 24%. Additionally, 18 out of 29 have participated in V3, and 6 out of 12 have participated in V4.

## 4. Discussion

One of modern medicine’s success stories is the development of personalized medicine approaches in oncology and immunology [68]. In oncology, for example, diagnostic and therapeutic decision-making is often guided by biomarker analyses of tumor samples and bodily fluid samples (e.g., liquid biopsy) [69]. This innovation has been achieved mostly through deep profiling of the biological underpinnings, providing critical insights into the pathophysiological factors contributing to these heterogeneous disorders [70].

Prognostic biomarkers in psychiatry could tremendously reduce mortality, suffering, and socioeconomic burden associated with mental health disorders. This is especially important in light of new emerging treatments with different mechanisms of action [71],[72]. Nevertheless, predicting treatment outcomes and prognosis remains an enduring challenge despite decades of progress in neuroscience [73]. This challenge arises not only from limited accessibility to the affected tissue but also from the unique complexity of the psychosocial, anatomical, molecular, and genetic architectures of mental disorders combined with high interindividual heterogeneity [70]. We argue that just as in oncology, precision medicine in psychiatry should go hand in hand with deep multi-system profiling of the biological underpinnings, utilizing technological advances such as multi-omics, neuroimaging, and neurophysiology, along with deep clinical phenotyping. Prediction models based on routine clinical data alone, without additional biological markers, have limited generalizability [28]. A systems biology approach [70] is thus needed to enable the integration of multidimensional biological and clinical data, to predict treatment outcomes, and to explore the mechanisms behind these trajectories.

Despite the sparsity of multimodal, deeply phenotyped cohorts in schizophrenia research, some prognostic biomarker candidates exist, such as FSA score [18], glial cell line-derived factor (GFNF) levels in the cerebrospinal fluid [74], extracellular vesicle-based biomarkers [75], and brain structure measurements [76]. Importantly, while some of these biomarker candidates have been internally validated, to the best of our knowledge, none of the available prognostic biomarker candidates have been validated in an independent cohort, questioning their generalizability [29]. To close the generalizability gap of biomarker research in SSDs, we urgently need longitudinal, naturalistic, deeply phenotyped cohorts, which could enable external validation of such biomarker candidates. Without this crucial step, it remains unclear if any of these biomarker candidates could be translated into clinical practice.

Biomarker research in schizophrenia should not solely be data-driven but needs to be reciprocally connected to mechanistic research. Most of the biomarker candidates identified to date are linked to specific biological pathways and mechanistic nodes that are considered to play critical roles in the pathophysiology of schizophrenia spectrum disorders [18],[74]. This integrative approach—combining pragmatic, translational biomarker studies with mechanistic research—has the potential to advance both disciplines mutually. Validation and testing of biomarker candidates can yield novel insights into the biological underpinnings of treatment outcomes, and these mechanistic insights, in turn, enable further refinement of biomarkers to capture the underlying pathophysiological pathways of SSDs more accurately [70].

Previous work from our group provides evidence that integrating multimodal data [30] can generate multi-level biological insights into the underlying pathophysiology of SSDs and establish links to clinically relevant outcomes [77]. Leveraging personalized disease models and deep phenotyping, we could demonstrate that genetically driven changes in neuronal gene expression and a resulting reduction in excitatory synaptic density *in vitro* are linked to alterations of brain structure, electrophysiology, and cognitive functioning *in vivo* [9]. Using the retina as a model of the central nervous system (CNS), we could demonstrate multimodal microstructural [10] and electrophysiological retinal alterations in individuals with SSDs that are associated with disease severity, individual polygenic burden [57], and linked to disturbed synapse biology [59]. In light of mounting evidence for lively brain-body interactions as contributing factors to the pathophysiology of SSDs, we investigated the blood-brain barrier (BBB) and the blood-cerebrospinal fluid barrier (BCB) in SSDs [78]. In the largest dynamic contrast-enhanced MRI study in SSD to date, we reported higher BBB leakage in individuals with SSDs compared to healthy controls in multiple brain regions implicated in the disorder [79]. Furthermore, we found that BCB disruption was associated with both dyslipidemia and a history of clozapine treatment in SSDs [80]. The volumes of the choroid plexus, a central hub of the blood-CSF interface, showed higher variability in individuals with SSDs compared to healthy controls [3] and were positively associated with brain region volumes previously linked to peripheral inflammation [81].

Building on this prior work, the CDP-STAR study employs a longitudinal approach utilizing comprehensive, multimodal protocols. To address the replication crisis in psychiatry [29],[28], we specifically aim to 1) externally validate previously identified prognostic biomarker candidates. Additionally, we aim to 2) investigate the biological underpinnings of treatment outcomes in SSDs and 3) identify novel biomarker candidates and mechanistic targets to enrich our understanding of the underlying pathophysiology.

In summary, this approach will help to pave the way for more reliable and mechanistically informed biomarker-based stratification in psychiatry, ultimately improving patient care and long-term outcomes.

## Contributions

VY, EB, LR, and JM designed and conceptualized the CDP-STAR Study. VY, EB, LR, and JM wrote the protocol. VY, JM, EB, LN, MW, and MB recruited patients and collected study data. LR, DK, and BS provided supervision. VY wrote the first draft of the manuscript, and all authors commented on previous versions of the manuscript. All authors read and approved the final manuscript.

## Data Availability

All data produced in the present study are available upon reasonable request to the authors.

